# Landscape of non-SARS-CoV-2 respiratory virus sequence data in Africa

**DOI:** 10.64898/2026.06.27.26356737

**Authors:** Kirsty Kwok, Laura Mojsiejczuk, Joseph Hughes, Ana da Silva Filipe, Antonia Ho

## Abstract

**Background:** Sequencing capacity in Africa greatly expanded during the COVID-19 pandemic. However, the availability of sequence data for non-SARS-CoV-2 respiratory viruses in the region remains uncertain. We systematically analysed sequence data from GenBank, GISAID and Pathoplexus for 12 non-SARS-CoV-2 respiratory virus groups from Africa and compared Africa’s genomic record availability with global genomic datasets. We further examined global genomic sequences to identify virus-specific patterns that could inform respiratory virus monitoring efforts in Africa.

**Results:** In Africa, the most sequenced virus was respiratory syncytial virus (RSV) (n=15,452), followed by influenza A virus (IAV) (n=10,900) and rhinovirus (n=4,774), when all sequences, including partial ones, were considered. Kenya and South Africa together contributed more than 60% of African respiratory virus sequences, while 30% of African countries submitted none. Within the global dataset of near-complete genomes, IAV was the most sequenced virus in Africa, consistent with global trends. However, Africa contributed the second-lowest number of genomes per million population among all world regions, exceeding only Asia. Nonetheless, an overall steady improvement in genome coverage was observed in respiratory virus sequences from Africa over time.

Substantial differences in relative genetic diversity were observed across viruses. Cytomegalovirus (94%) and parechovirus (38%) showed high proportions of representative sequences after clustering, with their clusters typically comprising sequences from a single region. In contrast, influenza viruses and RSV exhibited lower proportions of representative sequences, with most clusters circulating across multiple world regions.

**Conclusions:** Our analysis revealed substantial regional disparities in respiratory virus sequence availability across Africa, along with differences in diversity and geographical distribution patterns observed among viruses. This indicates the need for tailored and virus-specific surveillance strategies. Expanding sequencing capacity in more African nations is crucial for a clearer understanding of respiratory viruses circulating in Africa. Ultimately, this can guide vaccine and diagnostic development as well as performance assessment, aiding pandemic preparedness through the timely detection of emerging respiratory viruses.

## Introduction

Acute respiratory infections (ARI) impose a significant disease burden worldwide. In 2021, an estimated 12.8 billion episodes of upper respiratory infection (URI) and 344 million episodes of lower respiratory infection (LRI), both excluding coronavirus disease 2019 (COVID-19), were recorded across all age groups globally [1, 2]. LRI is also the second leading cause of death among communicable diseases, surpassed only by COVID-19 [3], while URI contributes significantly to overall disease burden and healthcare utilisation [1]. Mortality rates associated with both non-COVID-19 URI and LRI are highest in sub-Saharan Africa, reflecting regional disparities in health outcomes [1, 2]. This is likely due to a combination of a high prevalence of comorbidities, such as human immunodeficiency virus (HIV) infection and undernutrition, as well as socioeconomic factors including poverty and limited access to quality healthcare in parts of the region [4, 5].

Over the past two decades, the emergence of novel respiratory viruses, including severe acute respiratory syndrome coronavirus (SARS-CoV), the 2009 pandemic influenza A (H1N1) virus, Middle East respiratory syndrome coronavirus (MERS-CoV), and severe acute respiratory syndrome coronavirus 2 (SARS-CoV-2), has highlighted the critical role of viruses in the aetiology of ARI [6]. In addition to these emerging viruses, endemic respiratory viruses such as respiratory syncytial virus (RSV), rhinovirus, human parainfluenza virus (HPIV), adenovirus, human metapneumovirus (HMPV) and endemic human coronavirus (HCoV), continue to contribute substantially to global ARI burden [7, 8], including Africa [4, 9].

The COVID-19 pandemic drove global investment in genomic sequencing, including the prompt expansion of sequencing capacity in Africa [10, 11]. Despite this, marked global disparities were noted in SARS-CoV-2 genomic surveillance, with under-representation from low- and middle-income countries, particularly in parts of Africa [12, 13]. Advances in genomic sequencing have facilitated the rapid detection of variants of concern, tracking virus transmission, adapting public health measures, and selecting SARS-CoV-2 vaccine strains during the pandemic [14–16]. These efforts underscored the vital importance of coordinated and large-scale sequencing initiatives in improving our understanding of the transmission and evolutionary patterns of respiratory viruses.

While SARS-CoV-2 sequencing efforts have been widely evaluated, the availability and characteristics of sequence data for other clinically important non-SARS-CoV-2 respiratory viruses remain poorly characterised. Addressing this gap is essential for informing broader surveillance strategies beyond pandemic contexts.

In this study, we systematically characterised sequence data for 12 non-SARS-CoV-2 respiratory viruses from Africa, using data retrieved from three major sequence repositories: GenBank, the Global Initiative on Sharing All Influenza Data (GISAID), and Pathoplexus. We further compared African near-complete genome data with global datasets to contextualise representation and identify disparities in sequencing efforts. Lastly, we analysed global genomic sequences to identify virus-specific patterns that may inform surveillance strategies tailored to the African context. Collectively, this work provides a comprehensive overview of the geographical distribution and diversity of non-SARS-CoV-2 respiratory virus sequence records in Africa, highlighting key strengths, gaps, and opportunities for strengthening genomic surveillance. Enhanced understanding of the genomic epidemiology of respiratory viruses in Africa has the potential to improve ARI management by informing diagnostic development and treatment guidelines, guiding vaccine strain selection, and supporting the response to emerging and re-emerging viruses.

## Methods

### Data collection

All available nucleotide sequence records of 12 study-defined groups of endemic human non-SARS-CoV-2 respiratory viruses, including influenza A (IAV), B (IBV) and C (ICV) viruses, RSV, HPIV 1–4, HCoV HKU1, OC43, NL63 and 229E, HMPV, rhinovirus, parechovirus, adenovirus, bocavirus, and cytomegalovirus (CMV), were retrieved from 3 sequence repositories: GenBank via [the NCBI Virus](https://www.ncbi.nlm.nih.gov/labs/virus/vssi/#/) portal, GISAID (IAV, IBV, ICV and RSV) and Pathoplexus (RSV and HMPV) between February and June 2026. GenBank sequences were identified based on their respective NCBI taxonomic identifiers (TaxId) (**Supplementary Table 1**) (**Figure 1**). IAV, IBV and ICV sequence data from GISAID were retrieved from EpiFlu™, and RSV data from EpiRSV™ [17]. RSV and HMPV sequence data from Pathoplexus were retrieved for each virus from the corresponding databases (RSV-A, RSV-B, and HMPV) [18]. Sequences with restricted access (embargoed sequences) from GISAID and Pathoplexus were excluded. The detailed search strategy and data curation process are described in **Supplementary Materials**. Our analysis comprised two primary datasets: 1) the African dataset, including all available partial and complete sequences collected from the continent, and 2) the global genomic dataset restricted to high-quality sequences with ≥90% genome coverage.

**Figure 1.**
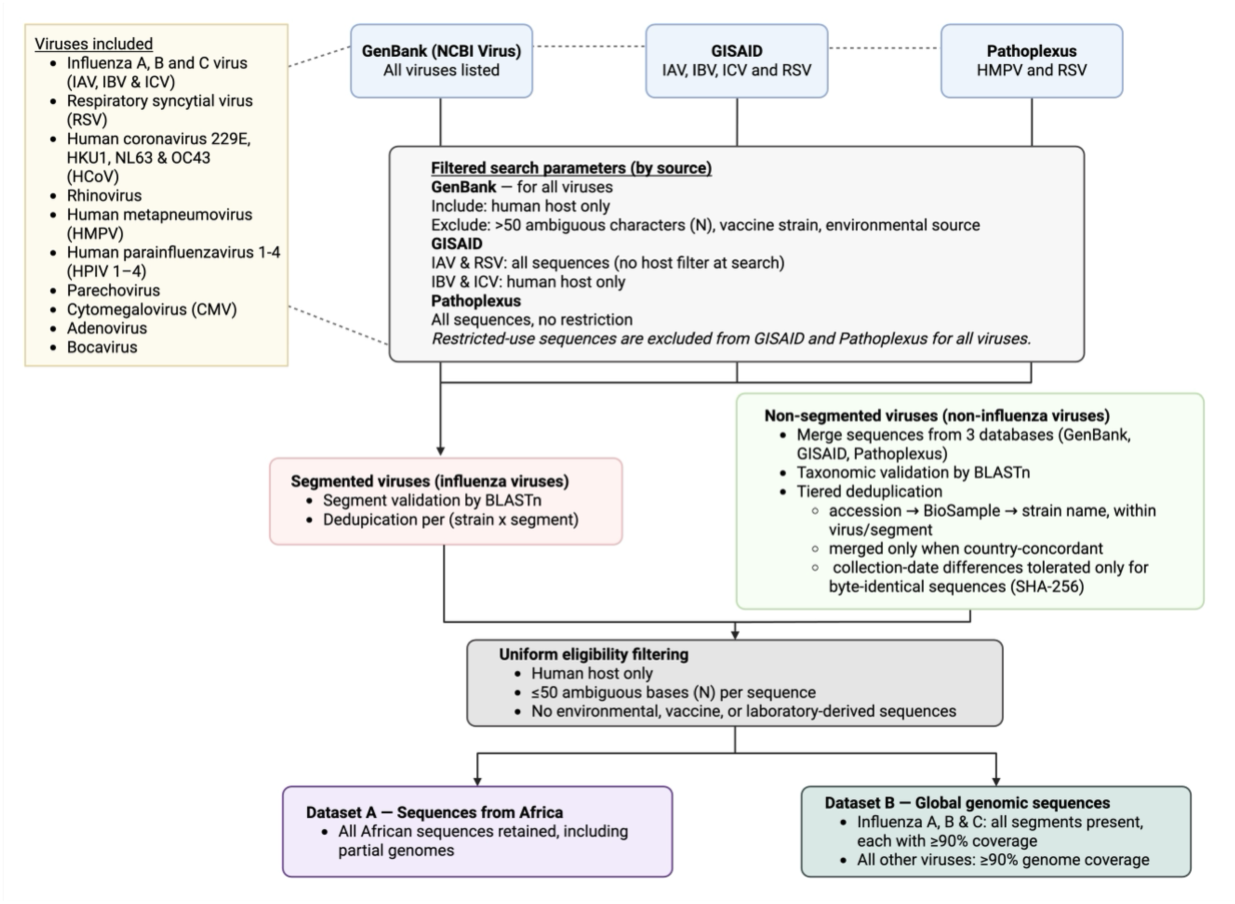
Flow diagram showing the sequence record curation process for the African sequence dataset (purple) and global genomic sequence dataset (green).

### Data analysis

All sequences with segment/genome coverage of ≥ 90% underwent nucleotide clustering using MMseqs2[19] with a 95% sequence identity threshold to compare the relative genetic diversity across different respiratory viruses. Summary statistics and data visualisation were performed using R packages and biorender.com (**Supplementary Table 2**).

## Results

### Distribution of non-SARS-CoV-2 respiratory virus sequences in Africa

A total of 37,872 deduplicated non-SARS-CoV-2 respiratory virus sequence records from Africa were included in this study (**Supplementary Table 3**). Over half of the sequences were obtained from GISAID (53%, n=20,404), which, at the time of analysis, only hosted IAV, IBV, ICV and RSV sequences. GenBank contributed 43% (n=16,333), and was the one repository covering all respiratory viruses examined in this study. Pathoplexus accounted for the remaining 3%, comprising RSV and HMPV sequences. The earliest sequence in this dataset was an IBV from South Africa, collected in 1958.

RSV (n=15,452) and IAV (n=10,900) were the most frequently sequenced viruses, followed by rhinovirus (n=4,774), IBV (n=4,368), and adenovirus (n=665) (**Supplementary Table 3**). IAV sequences were reported by 35 African countries, representing the highest geographical coverage, followed closely by IBV (34 countries). In contrast, RSV and rhinovirus sequences were only available in 22 and 12 countries, respectively, while ICV had the lowest representation, with only 4 sequences.

All five African sub-regions contributed sequence data for RSV, IAV, rhinovirus, IBV, adenovirus, HMPV, bocavirus, and CMV, with Eastern Africa contributing the largest share for most viruses. An exception was bocavirus, where Northern Africa predominated (**Supplementary Figure 1**). In contrast, HCoV and parechovirus sequences were reported from only three sub-regions (Eastern, Middle, and Western Africa), while HPIV and ICV were limited to two sub-regions each.

Kenya contributed the largest proportion of sequences (39%, n=14,798), followed by South Africa (21%, n=8,298) (**Figure 2**). Notably, 30% (n=16) of African countries had no respiratory virus sequences. Among the 38 countries/territories with available data (excluding Kenya and South Africa to minimise skew), 13% (n=5) contributed 1,000-1,500 sequences, 50% (n=19) contributed 100–999 sequences, 26 (n=10) submitted 10–99, and 11% (n=4) had fewer than 10 sequences.

**Figure 2.**
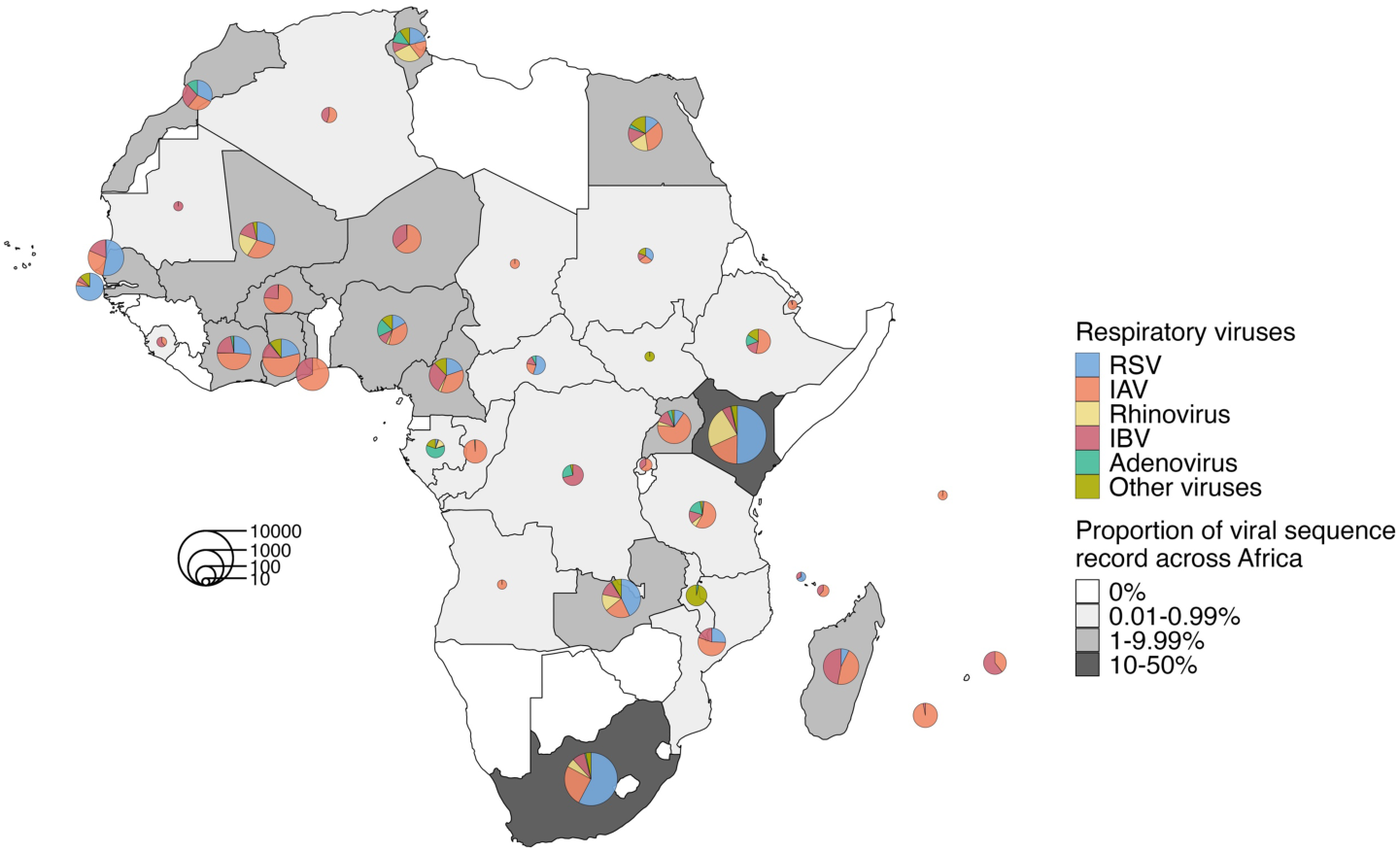
Distribution of respiratory virus sequences across Africa. Scatterpie charts are scaled to the number of deduplicated sequence records per country/territory, with slices showing the proportion of the five most sequenced viruses (RSV, IAV, rhinovirus, IBV and adenovirus) and other viruses, including HMPV, ICV, HPIV, HCoV, CMV, bocavirus, and parechovirus, grouped. Grayscale shading indicates each country’s share of the total number of African sequences.

Regarding viral diversity, 30% of contributing countries reported sequence data for more than five virus groups, with South Africa (10 virus groups), and Kenya (9 virus groups) showing the greatest diversity. In contrast, 58% of countries reported data for two and five virus groups, and 13% reported one single virus group.

To assess potential geographical structuring, nucleotide clustering at 95% identity was performed on near-complete genomes with genome coverage ≥90%. No clear associations were observed between geographical proximity and cluster-sharing frequency (**Supplementary Figure 2A**–**C**). Instead, higher proportions of shared clusters between countries correlated with greater sequencing output, as reflected by the number of sequences contributed per country (**Supplementary Figure 2D**–**E**).

### Sequencing strategies and data reporting characteristics

More than 60% of countries/territories (n=25) had sequences submitted by in-country organisations, used here as a proxy for local sequencing capacity. However, 45% of countries/territories had the majority of their sequences submitted by out-of-country organisations, particularly in Eastern, Western and Middle Africa (**Figure 3A**).

**Figure 3.**
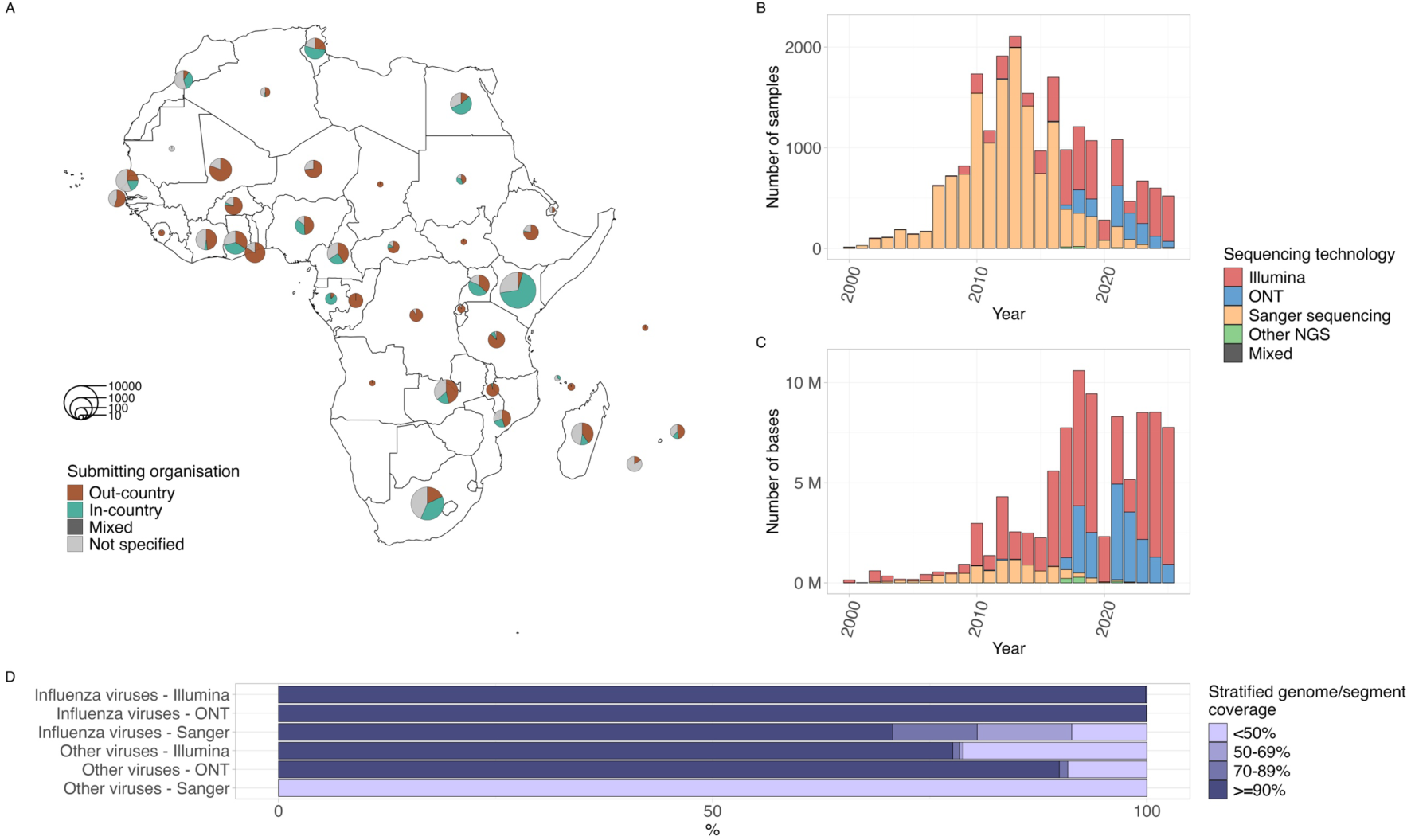
Overview of sequencing strategies used to generate respiratory virus sequences from Africa. (A) Distribution of the origin of submitting organisations for non-SARS-CoV-2 respiratory virus sequences across Africa. Scatterpie charts are scaled to the number of deduplicated sequence records per country/territory, with slices showing the proportion of submissions by each origin category. (B) Total number of deduplicated samples and (C) total number of bases submitted per collection year for respiratory virus sequences from Africa. Bars are coloured by the sequencing technology used. (D) Distribution of stratified segment coverage of influenza viruses and genome coverage of other viruses obtained using different sequencing platforms. The X-axis indicates the proportion of deduplicated sequence records.

Sequencing technology metadata were available for most GenBank records and for RSV sequences on GISAID, but not for influenza viruses on GISAID. In Pathoplexus, the field exists but was left blank for all records. Consequently, sequences from GISAID and Pathoplexus were excluded from this analysis. Among GenBank records, sequences from eight countries were generated exclusively using Sanger sequencing (**Supplementary Figure 3**). In contrast, Illumina sequencing was reported in 17 countries, while Oxford Nanopore Technology (ONT) was used in only four countries (Kenya, Cameroon, Uganda and Tanzania).

Sequencing output has steadily risen since 2000, peaking in 2012, followed by fluctuations in subsequent years. A marked decline was observed in 2020, with moderate recovery thereafter (**Figure 3B**). Prior to 2017, most sequences were generated using Sanger sequencing, whereas a more balanced use of different platforms was observed from 2017 onwards, with increasing adoption of next-generation sequencing (NGS) technologies, including Illumina and ONT. NGS platforms were associated with higher sequencing output (**Figure 3C**) and improved genome coverage, especially for non-influenza viruses, compared to Sanger-derived sequences (**Figure 3D**).

For influenza viruses, segment coverage improved substantially from 2006 onwards (**Supplementary Figure 4**), with segment coverage ≥90% achieved in 90% of sequences by 2011. In contrast, for other viruses, the proportion of sequences with ≥90% genome coverage first reached 50% only in 2019, declined briefly in 2021, and subsequently rose again, surpassing 90% by 2024.

Reporting turnaround time (TAT), defined as the interval between sample collection and sequence release, varied widely over time (**Supplementary Figure 5A**). The median TAT peaked in 2021 at over 3,000 days and has since shortened thereafter. Sequences with a rapid TAT (<21 days) were first observed in 2024. Across viruses, four had a median TAT below one year, with IAV the shortest (105 days), followed by IBV (150 days), ICV (202 days), and CMV (221 days) (**Supplementary Figure 5B**). However, only IAV, IBV and RSV had sequences with a TAT under 21 days.

Metadata completeness for specimen type was limited, with 86% of sequences lacking this information. Among those with available data, most viruses were predominantly associated with respiratory specimens. Exceptions included adenovirus, bocavirus, and parechovirus, which were more frequently detected in faecal samples, and CMV, which was more commonly associated with blood samples (**Supplementary Figure 6**).

### African genomic data in the global context

To contextualise Africa’s sequencing activity globally, we analysed the distribution of respiratory virus sequences with ≥90% genome coverage across all regions (**Figure 1**). A total of 316,370 deduplicated genome records were identified (**Supplementary Table 4**). IAV was the most frequently sequenced virus, comprising 66.2% of all records (n=209,461), followed by IBV (17.0%, n=53,679) and RSV (13.2%, n=41,781). ICV (n=260) and CMV (n=308) were the least represented viruses.

By repository, GISAID contributed nearly two-thirds of sequences (66.1%, n= 208,966), followed by GenBank (33.7%, n= 106,692), and Pathoplexus (0.2%, n=712) in the near-complete genome dataset. When normalised by population size, Asia (8.9 sequences per million population) and Africa (9.2 sequences per million population) had the lowest sequencing output among regions, whereas Oceania (408 sequences per million population) submitted the most (**Figure 4**). Across all regions, IAV, IBV, and RSV were the three most frequently sequenced viruses. Although RSV was the most-sequenced virus in Africa when partial sequences were included, IAV accounted for the largest proportion of near-complete genomes.

**Figure 4.**
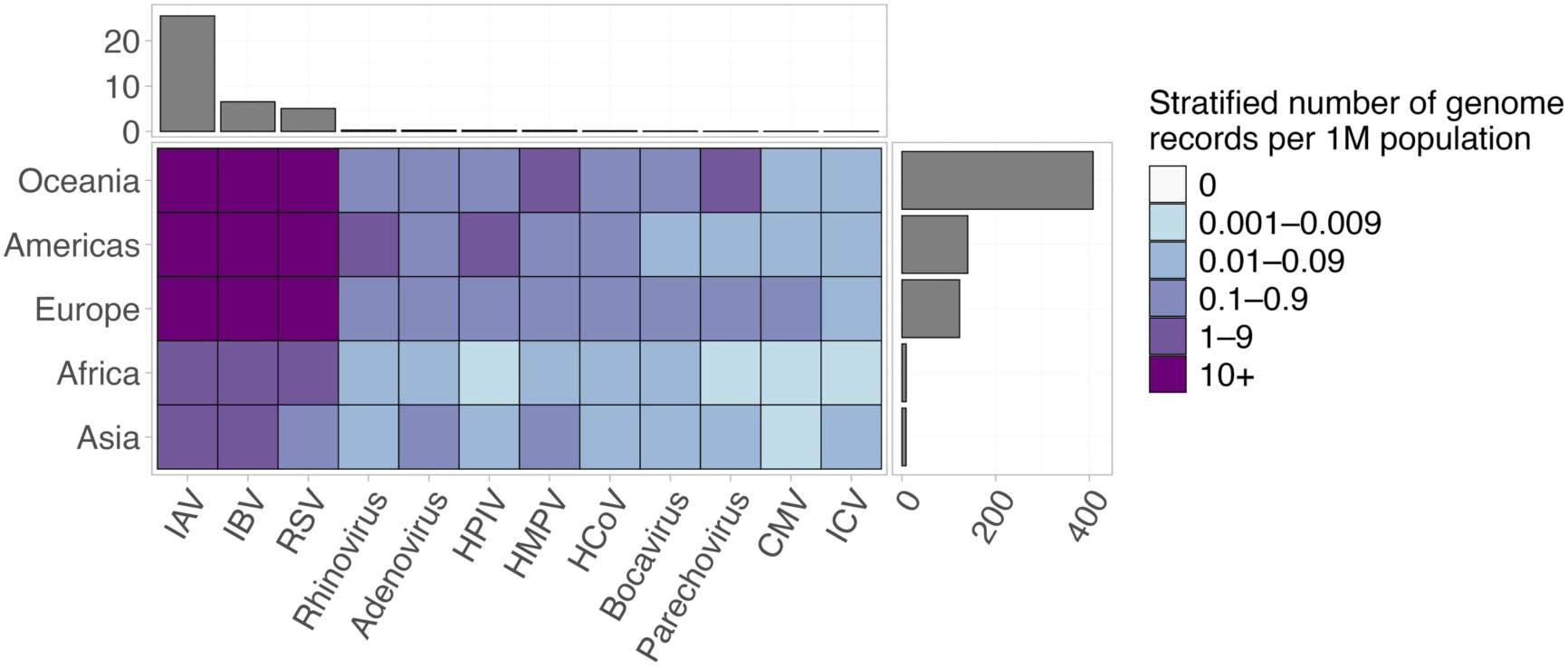
Global distribution of respiratory virus genomic sequences. The cell colour of the heatmap shows the stratified number of deduplicated genome sequence records per million population by virus and region. Bar plots indicate the total number of deduplicated genome sequence records per million population per virus (top) and per region (right).

Longitudinal analysis showed consistent sequencing activity over time for IAV, IBV, and RSV across all regions, with a noticeable dip between 2020 and 2021 followed by a gradual recovery from 2022 onwards (**Supplementary Figure 7**). Conversely, sequencing submissions for other viruses were comparatively limited and sporadic across both time and regions. The number of years with at least one available sequence for these viruses (i.e. excluding IAV, IBV and RSV) differed significantly between regions (Friedman’s test, p-value <0.0001, Kendall’s W = 0.8), with Oceania and Africa showing the shortest temporal coverage (medians of 5 and 6 years, respectively).

Within Africa, the temporal coverage varied considerably across viruses. Adenovirus sequences spanned 12 distinct years, whereas ICV was represented in only one single year. Intermediate temporal coverage was observed for HMPV and bocavirus sequences (9 years), rhinovirus (7 years), HCoV (6 years), HPIV (5 years), parechovirus (4 years), and CMV (3 years).

### Global comparative genomic and temporal patterns of non-SARS-CoV-2 respiratory viruses

To complement the limited genomic data from Africa across all 12 non-SARS-CoV-2 respiratory viruses, we analysed global datasets to characterise virus-specific patterns in genetic diversity and geographical distribution. Nucleotide clustering revealed substantial variation in the proportion of representative sequences across viruses (**Figure 5A**). IBV (0.01%), IAV (0.05%), and RSV (0.3%), the viruses with the largest datasets, exhibited the lowest proportion of representative sequences after clustering at 95% nucleotide identity. In contrast, CMV showed the highest retention (94%), followed by parechovirus (38%) and rhinovirus (12%), suggesting greater relative genetic diversity within available genomes.

**Figure 5.**
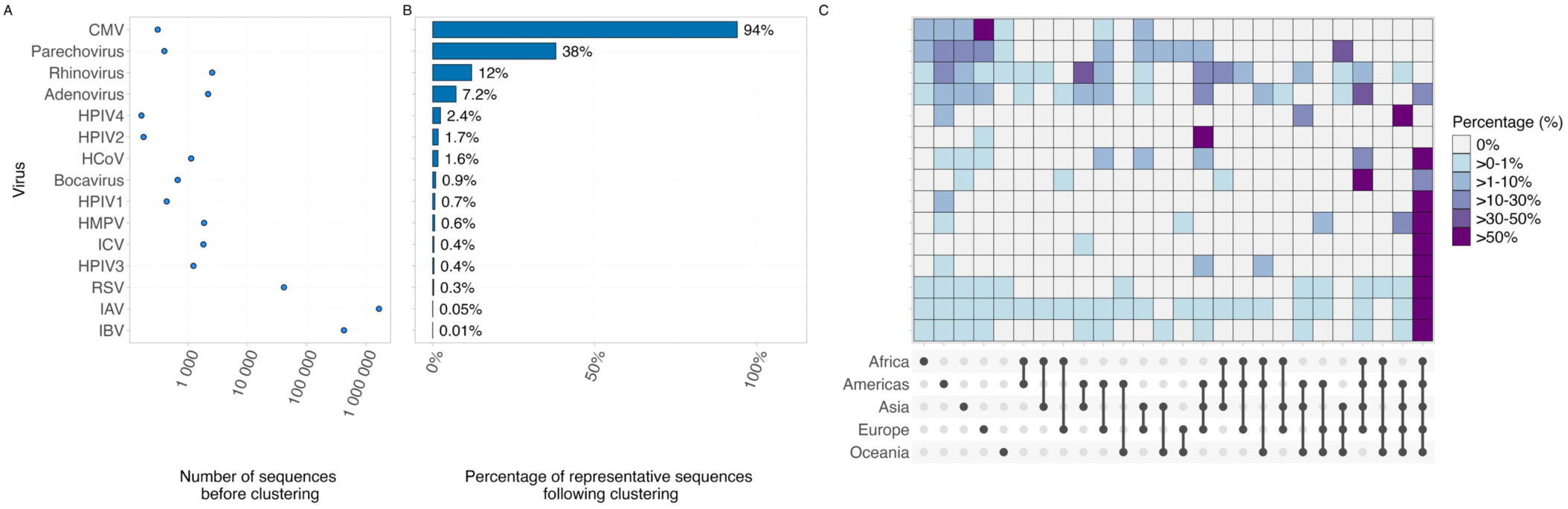
Overview of sequence diversity and interregional sharing of representative viral clusters (A) Number of sequences of each virus before MMSeqs2 clustering (x-axis shown on a log scale). (B) Percentage of representative sequences of each virus after MMSeqs2 clustering. (C) Regional sharing of representative sequence clusters by virus. The colour of each cell shows the percentage of clusters containing sequences from specific combinations of regions by virus, based on MMSeqs2 clustering and sequence geographical origins. The bottom node bar denotes the geographical region(s) represented within each representative sequence cluster.

We next examined the geographical composition of multi-sequence clusters to explore spatial patterns (**Figure 5B**). CMV (98%) and parechovirus (52%) exhibited a higher proportion of region-specific clusters, suggesting more localised transmission. Rhinovirus showed geographical structuring, with 19% of clusters confined to a single region and 42% spanning two regions. Meanwhile, several other viruses, including IAV, IBV, ICV, HPIV1, HPIV3, and RSV, displayed proportions of geographically widespread clusters exceeding 95%, suggesting extensive global dissemination. Similarly, a higher proportion of clusters for HCoV (85%) and HMPV (70%) included sequences from all five regions, consistent with widespread global circulation.

Lastly, we compared the temporal persistence of viral clusters by measuring the interval between the earliest and most recent sequences within each non-singleton cluster (**Supplementary Figure 8**). Substantial heterogeneity in persistence was noted. ICV exhibited the longest median cluster duration (74 years), followed by HPIV1 (44 years), IBV (31 years), HMPV (31 years), bocavirus (19 years), adenovirus (18 years), HPIV2 (16 years), HPIV4 (16 years), HCoV (15 years), and HPIV3 (14 years). Short persistence was observed for IAV and rhinovirus (6 years each), CMV (2 years), parechovirus (1 year), and RSV (0 years). However, estimates for bocavirus and HPIV 1–4 should be interpreted with caution due to the limited number of clusters (n ≤ 5).

## Discussion

In this study, we systematically characterised the geographical distribution, genetic diversity, and sequencing strategies of sequence records from three major sequence repositories for 12 non-SARS-CoV-2 respiratory viruses in Africa. This region was notably under-represented in genomic surveillance during the early stages of the COVID-19 pandemic [12]. Nevertheless, SARS-CoV-2 sequence output in Africa (185,392 sequences on GISAID as of 21^st^ June 2026) still exceeds that of all other respiratory viruses combined, reflecting the scale of global investment during the pandemic.

When considering all sequences, including both complete and partial, RSV was the most sequenced non-SARS-CoV-2 respiratory virus in Africa, followed by rhinovirus and IAV. This distribution likely reflects the research priorities in the region, supported by multiple cohort and retrospective studies focusing on RSV and rhinovirus epidemiology and evolution in sub-Saharan Africa [20–24]. However, when restricting analysis to near-complete genomes, IAV was the most frequently sequenced, consistent with global trends. This pattern likely reflects sustained sequencing efforts through the World Health Organisation (WHO) Global Influenza Surveillance and Response System (GISRS), a well-established international surveillance programme [25].

Despite the disproportionate burden of ARI in Africa, it ranked among the lowest regions globally for respiratory virus genomic sequences per capita (9.2 sequences per million population), slightly above Asia (8.9 sequences per million population). This disparity possibly reflects differences in sequencing capacity and surveillance priorities, with many African countries allocating their limited resources towards high-burden diseases, such as HIV, malaria and tuberculosis [26, 27]. Furthermore, within Africa, non-SARS-CoV-2 respiratory virus sequence data were highly unevenly distributed. Kenya and South Africa together accounted for more than 60% of all sequences, with Kenya alone contributing nearly 40%. These findings were consistent with previous reports identifying these countries as having the highest genome sequencing capacity on the continent at the onset of the Africa Pathogen Genomics Initiative (PGI) [28]. This geographic concentration mirrors reports of heterogeneous SARS-CoV-2 genomic surveillance efforts in the region [12, 13], although the disparity was somewhat less pronounced in SARS-CoV-2 sequencing. This is likely due to targeted investments during the pandemic, with other countries such as The Gambia, Djibouti, and Burkina Faso sequencing a higher proportion of cases [12]. Initiatives such as the Africa PGI, launched by the Africa Centres for Disease Control and Prevention (Africa CDC) in late 2020 with substantial funding, played a key role in expanding sequencing capacity across the continent [28].

Despite this progress, 30% of African countries did not submit any respiratory virus sequences, and in many others, sequence submissions were dominated by out-of-country organisations. While over 60% of African countries/territories had some level of in-country sequence submission, external contributions still accounted for the majority of sequences in nearly half of the countries. This metric should be interpreted with caution, as the metadata may not fully capture collaborative arrangements, but it nevertheless highlights the ongoing need to strengthen local sequencing capacity and ownership.

Encouragingly, our analysis showed that sequences generated using NGS platforms were generally more complete than those obtained through Sanger sequencing, with clear improvements in genome and segment coverage over time. This trend aligns with the increasing adoption of NGS technologies across the continent and underscores their importance for high-resolution genomic surveillance. Sustained investment in infrastructure, workforce development and supply chains will be essential to maintain and expand these gains.

Reported TAT differed substantially across viruses. Influenza viruses, particularly IAV and IBV, had the shortest median TATs (<5 months). This is likely due to GISAID’s inclusion of sequence records from WHO GISRS and other surveillance programmes, alongside its semi-restricted data-use model that encourages rapid sharing by allowing contributors to retain control and submit data prior to publication, thereby reducing TAT. Similarly, Pathoplexus enables time-limited restrictions (up to one year) before automatic release to the International Nucleotide Sequence Database Collaboration (INSDC) databases, balancing contributor recognition with eventual open access. Such models, which incentivise sharing while ensuring data openness, are critical to minimising reporting delays and enabling real-time monitoring of evolving respiratory viruses.

Our analysis did not identify a clear relationship between geographical proximity and sequence cluster sharing within Africa. Instead, cluster-sharing patterns appeared to correlate more strongly with sequencing intensity, suggesting that current datasets may be insufficient to resolve spatial transmission dynamics. The uneven distribution and limited volume of sequence data likely constrain such analyses, though factors such as human mobility and transportation networks may also contribute to viral spread across the region [29, 30]. Expanded and more geographically representative sequencing, combined with mobility data, will be critical to understand transmission dynamics.

At the global level, we observed marked heterogeneity in genetic diversity, geographical distribution, and temporal persistence across respiratory viruses. IAV, IBV, and RSV exhibited low proportions of representative sequences following clustering, indicating lower relative genetic diversity among available sequences. These viruses were also characterised by predominantly globally shared clusters, consistent with rapid international spread. Conversely, CMV, rhinovirus, and parechovirus retained more representative sequences and displayed more region-specific clustering, suggesting greater diversity and more localised transmission dynamics.

Temporal analysis further highlighted differences in lineage persistence. Viruses such as RSV, parechovirus, CMV, rhinovirus, and IAV showed short duration, indicative of rapid lineage turnover, whereas others (e.g. ICV and HPIVs) showed longer persistence. These differences emphasise that genomic surveillance strategies should be tailored to virus-specific characteristics. In resource-limited settings, including many African countries, prioritisation may be necessary. Viruses exhibiting higher genetic diversity, rapid turnover, or evidence of region-specific clustering, such as CMV, parechovirus, and rhinovirus, may require more extensive and locally-focused sampling, rather than reliance on global data[31]. Notably, genomic data for both CMV and parechovirus remain scarce in Africa, representing important gaps in current surveillance.

This study has several limitations. First, sparse sequence coverage and incomplete metadata, including missing specimen type information, constrained some analyses and underscored the need for improved adherence to the FAIR (Findable, Accessible, Interoperable, Reusable) principles [32] and the STROME-ID statement (Strengthening the Reporting of Molecular Epidemiology for Infectious Diseases) [33]. Additionally, geospatial analyses were limited to the country level due to the absence of subnational coordinates in the metadata. Second, the dataset reflects sequence data accumulated over time rather than systematic surveillance and may therefore be subject to sampling or reporting biases driven by research priorities, outbreak responses, or individual case studies. Third, while our MMseqs2-based clustering is well-suited to summarising relative genomic diversity across respiratory viruses in our dataset, future work could incorporate phylogenetic and phylodynamic analyses for resolving detailed evolutionary and transmission patterns. Despite these limitations, this analysis provides a comprehensive comparative overview of non-SARS-CoV-2 respiratory virus sequence data from Africa and places these findings in a global context. The results highlight both progress and persistent gaps in genomic surveillance across the continent.

Strengthening genomic surveillance in Africa remains critical, as evidenced by the identification of the Beta and Omicron SARS-CoV-2 variants in South Africa [14, 15], illustrating the potential for novel variant emergence, likely driven by persistent infections in immunocompromised populations in the region [34]. While recent investments driven by the COVID-19 pandemic and sporadic outbreaks of Ebola and mpox have improved public health sequencing capacity in Africa [11], challenges remain, including workforce training and retention, and reliable access to laboratory consumables [35]. Addressing these constraints will require sustained, coordinated, and collaborative efforts [36].

Cost-effective sequencing approaches, such as amplicon sequencing, may support continued surveillance of endemic respiratory viruses [31, 37], although these methods may be less suitable for highly diverse viruses. Regional capacity sharing, as demonstrated during the COVID-19 pandemic, may also pragmatically strengthen respiratory virus sequencing across the continent [10]. In the longer term, expanding and sustaining in-country genomic surveillance capacity across a broader range of respiratory pathogens and geographical locations will be essential to improve data representativeness and support more effective regional and global responses to ARI.

## Data availability

Sequences from GenBank are available on GenBank. Sequences from GISAID and Pathoplexus are available under dataset identifiers (GISAID: [EPI_SET_260622eb](https://epicov.org/epi3/epi_set/260622eb), [EPI_SET_260622ce](https://doi.org/10.55876/gis8.260622ce); Pathoplexus: PP_SS_2509.1), with corresponding DOI links listed in references [38–40].

## Contributions

Conceptualisation: All authors. Data curation: KK. Formal analysis: KK. Investigation: All authors. Writing: original draft: KK. Writing: reviewing & editing: All authors. Funding acquisition: JH, AdSF, AH. Validation: AdSF. Supervision: JH, AdSF, AH.

## Acknowledgements

We gratefully acknowledge all data contributors, i.e., the Authors and their Originating laboratories responsible for obtaining the specimens, and their Submitting laboratories for generating the genetic sequence and metadata and sharing via the GISAID Initiative, on which this research is based. Equally, we would like to thank all contributors who generated and submitted sequences via GenBank and Pathoplexus. We thank Christopher Illingworth and Rubayet Alam for helpful suggestions and advice.

## Ethics declarations

### Ethics approval and consent to participate

Not applicable.

### Consent for publication

Not applicable.

## References

1. Sirota SB, Doxey MC, Dominguez R-MV, Bender RG, Vongpradith A, Albertson SB, et al. Global, regional, and national burden of upper respiratory infections and otitis media, 1990–2021: a systematic analysis from the Global Burden of Disease Study 2021. The Lancet Infectious Diseases. 2025;25:36–51. 10.1016/S1473-3099(24)00430-4.

2. Bender RG, Sirota SB, Swetschinski LR, Dominguez R-MV, Novotney A, Wool EE, et al. Global, regional, and national incidence and mortality burden of non-COVID-19 lower respiratory infections and aetiologies, 1990–2021: a systematic analysis from the Global Burden of Disease Study 2021. The Lancet Infectious Diseases. 2024;24:974–1002. 10.1016/S1473-3099(24)00176-2.

3. World Health Organization. Global health estimates 2019: deaths by cause, age, sex, by country and by region, 2000-2019. 2020.

4. McMorrow ML, Wemakoy EO, Tshilobo JK, Emukule GO, Mott JA, Njuguna H, et al. Severe acute respiratory illness deaths in sub-Saharan Africa and the role of influenza: a case series from 8 countries. The Journal of Infectious Diseases. 2015;212:853–60. 10.1093/infdis/jiv100.

5. Sarfo JO, Amoadu M, Gyan TB, Osman A-G, Kordorwu PY, Adams AK, et al. Acute lower respiratory infections among children under five in Sub-Saharan Africa: a scoping review of prevalence and risk factors. BMC Pediatr. 2023;23:225. 10.1186/s12887-023-04033-x.

6. Gray GC, Abdelgadir A. While we endure this pandemic, what new respiratory virus threats are we missing? Open Forum Infect Dis. 2021;8:ofab078. 10.1093/ofid/ofab078.

7. Shi T, McLean K, Campbell H, Nair H. Aetiological role of common respiratory viruses in acute lower respiratory infections in children under five years: A systematic review and meta-analysis. J Glob Health. 2015;5:010408. 10.7189/jogh.05.010408.

8. Shi T, Arnott A, Semogas I, Falsey AR, Openshaw P, Wedzicha JA, et al. The etiological role of common respiratory viruses in acute respiratory infections in older adults: a systematic review and meta-analysis. The Journal of Infectious Diseases. 2020;222 Supplement_7:S563–9. 10.1093/infdis/jiy662.

9. Kenmoe S, Tchendjou P, Vernet M-A, Moyo-Tetang S, Mossus T, Njankouo-Ripa M, et al. Viral etiology of severe acute respiratory infections in hospitalized children in Cameroon, 2011–2013. Influenza and Other Respiratory Viruses. 2016;10:386–93. 10.1111/irv.12391.

10. Tegally H, San JE, Cotten M, Moir M, Tegomoh B, Mboowa G, et al. The evolving SARS-CoV-2 epidemic in Africa: Insights from rapidly expanding genomic surveillance. Science. 2022;378:eabq5358. 10.1126/science.abq5358.

11. Hill V, Githinji G, Vogels CBF, Bento AI, Chaguza C, Carrington CVF, et al. Toward a global virus genomic surveillance network. Cell Host & Microbe. 2023;31:861–73. 10.1016/j.chom.2023.03.003.

12. Chen Z, Azman AS, Chen X, Zou J, Tian Y, Sun R, et al. Global landscape of SARS-CoV-2 genomic surveillance and data sharing. Nat Genet. 2022;54:499–507. 10.1038/s41588-022-01033-y.

13. Brito AF, Semenova E, Dudas G, Hassler GW, Kalinich CC, Kraemer MUG, et al. Global disparities in SARS-CoV-2 genomic surveillance. Nat Commun. 2022;13:7003. 10.1038/s41467-022-33713-y.

14. Tegally H, Wilkinson E, Giovanetti M, Iranzadeh A, Fonseca V, Giandhari J, et al. Detection of a SARS-CoV-2 variant of concern in South Africa. Nature. 2021;592:438–43. 10.1038/s41586-021-03402-9.

15. Viana R, Moyo S, Amoako DG, Tegally H, Scheepers C, Althaus CL, et al. Rapid epidemic expansion of the SARS-CoV-2 Omicron variant in southern Africa. Nature. 2022;603:679–86. 10.1038/s41586-022-04411-y.

16. da Silva Filipe A, Shepherd JG, Williams T, Hughes J, Aranday-Cortes E, Asamaphan P, et al. Genomic epidemiology reveals multiple introductions of SARS-CoV-2 from mainland Europe into Scotland. Nat Microbiol. 2021;6:112–22. 10.1038/s41564-020-00838-z.

17. Elbe S, Buckland-Merrett G. Data, disease and diplomacy: GISAID’s innovative contribution to global health. Global Challenges. 2017;1:33–46. 10.1002/gch2.1018.

18. Vecchia ED. Pathoplexus: towards fair and transparent sequence sharing. The Lancet Microbe. 2024;5. 10.1016/j.lanmic.2024.100995.

19. Steinegger M, Söding J. MMseqs2 enables sensitive protein sequence searching for the analysis of massive data sets. Nat Biotechnol. 2017;35:1026–8. 10.1038/nbt.3988.

20. Smuts HE, Workman LJ, Zar HJ. Human rhinovirus infection in young African children with acute wheezing. BMC Infectious Diseases. 2011;11:65. 10.1186/1471-2334-11-65.

21. Agoti CN, Otieno JR, Munywoki PK, Mwihuri AG, Cane PA, Nokes DJ, et al. Local evolutionary patterns of human respiratory syncytial virus derived from whole-genome sequencing. Journal of Virology. 2015;89:3444–54. 10.1128/jvi.03391-14.

22. Baillie VL, Moore DP, Mathunjwa A, Morailane P, Simões EAF, Madhi SA. Molecular subtyping of human rhinovirus in children from three Sub-Saharan African countries. Journal of Clinical Microbiology. 2019;57:10.1128/jcm.00723-19. https://doi.org/10.1128/jcm.00723-19.

23. Ihling CM, Schnitzler P, Heinrich N, Mangu C, Sudi L, Souares A, et al. Molecular epidemiology of respiratory syncytial virus in children in sub-Saharan Africa. Tropical Medicine C International Health. 2021;26:810–22. 10.1111/tmi.13573.

24. Morobe JM, Kamau E, Luka MM, Murunga N, Lewa C, Mutunga M, et al. Spatio-temporal distribution of rhinovirus types in Kenya: a retrospective analysis, 2014. Sci Rep. 2024;14:22298. 10.1038/s41598-024-73856-0.

25. Steffen C, Debellut F, Gessner B, Kasolo F, Yahaya A, Ayebazibwe N, et al. Improving influenza surveillance in sub-Saharan Africa. Bull World Health Organ. 2012;90:301–5. 10.2471/BLT.11.098244.

26. Chola M, Sikazwe I, Robalo M, Oduro-Bonsrah P, Coutinho A, Sheneberger R, et al. Africa’s defining moment: the time to lead the HIV response is now. The Lancet Global Health. 2025;13:e801–2. 10.1016/S2214-109X(25)00102-0.

27. Gouda HN, Charlson F, Sorsdahl K, Ahmadzada S, Ferrari AJ, Erskine H, et al. Burden of non-communicable diseases in sub-Saharan Africa, 1990–2017: results from the Global Burden of Disease Study 2017. The Lancet Global Health. 2019;7:e1375–87. 10.1016/S2214-109X(19)30374-2.

28. Makoni M. Africa’s $100-million Pathogen Genomics Initiative. The Lancet Microbe. 2020;1:e318. 10.1016/S2666-5247(20)30206-8.

29. Browne A, St-Onge Ahmad S, Beck CR, Nguyen-Van-Tam JS. The roles of transportation and transportation hubs in the propagation of influenza and coronaviruses: a systematic review. Journal of Travel Medicine. 2016;23:tav002. 10.1093/jtm/tav002.

30. Gartland N, Fishwick D, Coleman A, Davies K, Hartwig A, Johnson S, et al. Transmission and control of SARS-CoV-2 on ground public transport: A rapid review of the literature up to May 2021. Journal of Transport & Health. 2022;26:101356. 10.1016/j.jth.2022.101356.

31. Pronyk PM, de Alwis R, Rockett R, Basile K, Boucher YF, Pang V, et al. Advancing pathogen genomics in resource-limited settings. Cell Genomics. 2023;3:100443. 10.1016/j.xgen.2023.100443.

32. Wilkinson MD, Dumontier M, Aalbersberg IjJ, Appleton G, Axton M, Baak A, et al. The FAIR Guiding Principles for scientific data management and stewardship. Sci Data. 2016;3:160018. 10.1038/sdata.2016.18.

33. Field N, Cohen T, Struelens MJ, Palm D, Cookson B, Glynn JR, et al. Strengthening the reporting of molecular epidemiology for infectious diseases (STROME-ID): an extension of the STROBE statement. The Lancet Infectious Diseases. 2014;14:341–52. 10.1016/S1473-3099(13)70324-4.

34. Sigal A, Neher RA, Lessells RJ. The consequences of SARS-CoV-2 within-host persistence. Nat Rev Microbiol. 2024;:1–15. 10.1038/s41579-024-01125-y.

35. Adebamowo SN, Francis, Veronica, Tambo, Ernest, Diallo, Seybou H., Landouré, Guida, Nembaware,Victoria, et al. Implementation of genomics research in Africa: challenges and recommendations. Global Health Action. 2018;11:1419033. 10.1080/16549716.2017.1419033.

36. Onywera H, Mulder N, Kebede Y, Tessema SK. How to sustain a public-health genomics and bioinformatics workforce in Africa. Nat Med. 2025;:1–5. 10.1038/s41591-025-03720-9.

37. KEMRI. Africa joins global effort to monitor emerging pathogens via genomics. KEMRI. 2025. https://www.kemri.go.ke/africa-joins-global-effort-to-monitor-emerging-pathogens-via-genomics/. Accessed 18 June 2025.

38. GISAID. EPI_SET_260622eb. 2026. 10.55876/gis8.260622eb.

39. GISAID. EPI_SET_260622ce. 2026. 10.55876/gis8.260622ce.

40. Pathoplexus. PP_SS_2509.1. 2026. 10.62599/PP_SS_2509.1.

