## Supplementary Materials for "Landscape of non-SARS-CoV-2 respiratory virus sequence data in Africa"

Supplementary Table 1	2

Supplementary Table 2	3

Supplementary Table 3	4

Supplementary Table 4	5

Supplementary Figure 1	6

Supplementary Figure 2	7

Supplementary Figure 3	8

Supplementary Figure 4	9

Supplementary Figure 5	10

Supplementary Figure 6	11

Supplementary Figure 7	12

Supplementary Figure 8	13

Supplementary Methods	14

Supplementary Table 1. A list of NCBI taxonomic identifiers (Taxid) used to perform our systematic search on NCBI GenBank nucleotide sequence records via the NCBI Virus portal.

| **Study-defined virus group** | **Also known as** | **NCBI Taxid** | **Species** | **Genus** | **Family** |
| --- | --- | --- | --- | --- | --- |
| Human parainfluenza virus (HPIV) | Human parainfluenza virus 1 | 12730 | *Respirovirus laryngotracheitidis* | *Respirovirus* | *Paramyxoviridae* |
|  |  | 3049952 | *Respirovirus laryngotracheitidis* | *Respirovirus* | *Paramyxoviridae* |
|  | Human parainfluenza virus 2 | 3052557 | *Orthorubulavirus laryngotracheitidis* | *Orthorubulavirus* | *Paramyxoviridae* |
|  |  | 2560525 | *Orthorubulavirus laryngotracheitidis* | *Orthorubulavirus* | *Paramyxoviridae* |
|  | Human parainfluenza virus 3 | 11216 | *Respirovirus pneumoniae* | *Respirovirus* | *Paramyxoviridae* |
|  |  | 3049953 | *Respirovirus pneumoniae* | *Respirovirus* | *Paramyxoviridae* |
|  | Human parainfluenza virus 4 | 3052556 | *Orthorubulavirus hominis* | *Orthorubulavirus* | *Paramyxoviridae* |
|  |  | 2560526 | *Orthorubulavirus hominis* | *Orthorubulavirus* | *Paramyxoviridae* |
|  |  | 11224 | *Orthorubulavirus hominis* | *Orthorubulavirus* | *Paramyxoviridae* |
|  |  | 11226 | *Orthorubulavirus hominis* | *Orthorubulavirus* | *Paramyxoviridae* |
| Rhinovirus | Rhinovirus | 147711 | *Enterovirus alpharhino* | *Enterovirus* | *Picornaviridae* |
|  |  | 147712 | *Enterovirus betarhino* | *Enterovirus* | *Picornaviridae* |
|  |  | 463676 | *Enterovirus cerhino* | *Enterovirus* | *Picornaviridae* |
|  |  | 169066 | N/A | *Enterovirus* | *Picornaviridae* |
|  |  | 348531 | N/A | *Enterovirus* | *Picornaviridae* |
| Influenza A virus (IAV) | Influenza A virus | 2955291 | *Alphainfluenzavirus influenzae* | *Alphainfluenzavirus* | *Orthomyxoviridae* |
| Influenza B virus (IBV) | Influenza B virus | 2955465 | *Betainfluenzavirus influenzae* | *Betainfluenzavirus* | *Orthomyxoviridae* |
| Influenza C virus (ICV) | Influenza C virus | 2955935 | *Gammainfluenzavirus influenzae* | *Gammainfluenzavirus* | *Orthomyxoviridae* |
| Human coronavirus (HCoV) | Human coronavirus 229E | 11137 | *Alphacoronavirus chicagoense* | *Alphacoroanvirus* | *Coronaviridae* |
|  | Human coronavirus OC43 | 31631 | *Betacoronavirus gravedinis* | *Betacoronavirus* | *Coronaviridae* |
|  | Human coronavirus NL63 | 277944 | *Alphacoronavirus amsterdamense* | *Alphacoroanvirus* | *Coronaviridae* |
|  | Human coronavirus HKU1 | 290028 | *Betacoronavirus hongkongense* | *Betacoronavirus* | *Coronaviridae* |
| Human respiratory syncytial virus (RSV) | Human respiratory syncytial virus | 3049954 | *Orthopneumovirus hominis* | *Orthopneumovirus* | *Pneumoviridae* |
| Human metapneumovirus (HMPV) | Human metapneumovirus | 3048148 | *Metapneumovirus hominis* | *Metapneumovirus* | *Pneumoviridae* |
| Adenovirus | Adenovirus | 10509 | N/A | *Mastadenovirus* | *Adenoviridae* |
| Bocavirus | Bocavirus | 1507401 | N/A | *Bocaparvovirus* | *Parvoviridae* |
| Cytomegalovirus (CMV) | Cytomegalovirus | 3050295 | *Cytomegalovirus humanbeta5* | *Cytomegalovirus* | *Orthoherpesviridae* |
| Parechovirus | Parechovirus | 138954 | N/A | *Parechovirus* | *Picornaviridae* |

Supplementary Table 2. A list of R packages used for data analyses

| **R package name (version)** | **Use** |
| --- | --- |
| data.table (v1.16.4)  dplyr (v1.1.4)  reshape2 (v1.4.4)  stringr (v1.5.1) | Data wrangling |
| ComplexHeatmap (v2.20.0)  ggplot2 (v3.5.1)  ggside (v0.3.1)  ggstatsplot (v0.12.4)  ggtext (v0.1.2)  ggupset (v0.4.1)  ggh4x (v0.3.0)  glue (v1.8.0)  patchwork (v1.3.0)  scatterpie (v0.2.4)  sf (v1.0.19) | Data visualisation |
| Rnaturalearth (v1.0.1)  rnaturalearthdata (v1.0.0) | Map data access |
| Base R package (R version 4.4.2)  ggstatsplot (v0.12.4) | Statistical analyses |

Supplementary Table 3**.** Overview of deduplicated sequence records from Africa

| **Virus** | **No. of deduplicated sequence records**  **(no. of segment sequences)** | **GenBank** | **GISAID** **(IAV, IBV, ICV and RSV)** | **Pathoplexus (HMPV, RSV)** | **No. of  countries /territories** | **Collection period** |
| --- | --- | --- | --- | --- | --- | --- |
| RSV | 15,452 | 6,103 | 8,307 | 1,042 | 22 | 1997-2025 |
| IAV | 10,900  (71806) | 2,712 | 8,188 | / | 35 | 1967-2026 |
| Rhinovirus | 4,774 | 4,774 | / | / | 12 | 2004-2023 |
| IBV | 4,368  (22,924) | 462 | 3,906 | / | 34 | 1958-2026 |
| Adenovirus | 665 | 665 | / | / | 17 | 1968-2025 |
| HMPV | 632 | 539 | / | 93 | 10 | 2007-2024 |
| Parechovirus | 306 | 306 | / | / | 8 | 2002-2019 |
| Bocavirus | 291 | 291 | / | / | 8 | 2005-2023 |
| CMV | 195 | 195 | / | / | 6 | 2002-2023 |
| HCoV | 144 | 144 | / | / | 7 | 2008-2023 |
| HPIV | 141 | 141 | / | / | 3 | 2007-2021 |
| ICV | 4  (16) | 1 | 3 | / | 2 | 1966-2017 |
| Total | 37,872 | 16,333 | 20,404 | 1,135 | / | / |

Supplementary Table 4**.** Overview of deduplicated genome records worldwide

| **Virus** | **No. of deduplicated genomes  (no. of segment sequences)** | **GenBank** | **GISAID (IAV, IBV, ICV and RSV)** | **Pathoplexus (HMPV and RSV)** | **Collection period** |
| --- | --- | --- | --- | --- | --- |
| IAV | 209,461  (1,675,592) | 71,684 | 137,777 | / | 1918-2026 |
| IBV | 53,679  (429,408) | 14,238 | 39,441 | / | 1940-2026 |
| RSV | 41,781 | 9,499 | 31,613 | 669 | 1956-2026 |
| Rhinovirus | 2,562 | 2,562 | / | / | 1999-2025 |
| Adenovirus | 2,195 | 2,195 | / | / | 1953-2025 |
| HPIV | 2,015 | 2,015 | / | / | 1957-2025 |
| HMPV | 1,906 | 1,863 | / | 43 | 1982-2025 |
| HCoV | 1,133 | 1,133 | / | / | 1983-2025 |
| Bocavirus | 670 | 670 | / | / | 2001-2025 |
| Parechovirus | 400 | 400 | / | / | 1993-2025 |
| CMV | 308 | 308 | / | / | 1956-2024 |
| ICV | 260  (1,820) | 125 | 135 | / | 1950-2024 |
| Total | 316,370 | 106,692 | 208,966 | 712 | / |

Supplementary Figure 1**.** Proportion of each respiratory virus’ genomes contributed by the five UNSD African sub-regions (Northern, Western, Middle, Eastern, Southern), with the total genome count per virus shown at the right. The top bar pools all viruses.

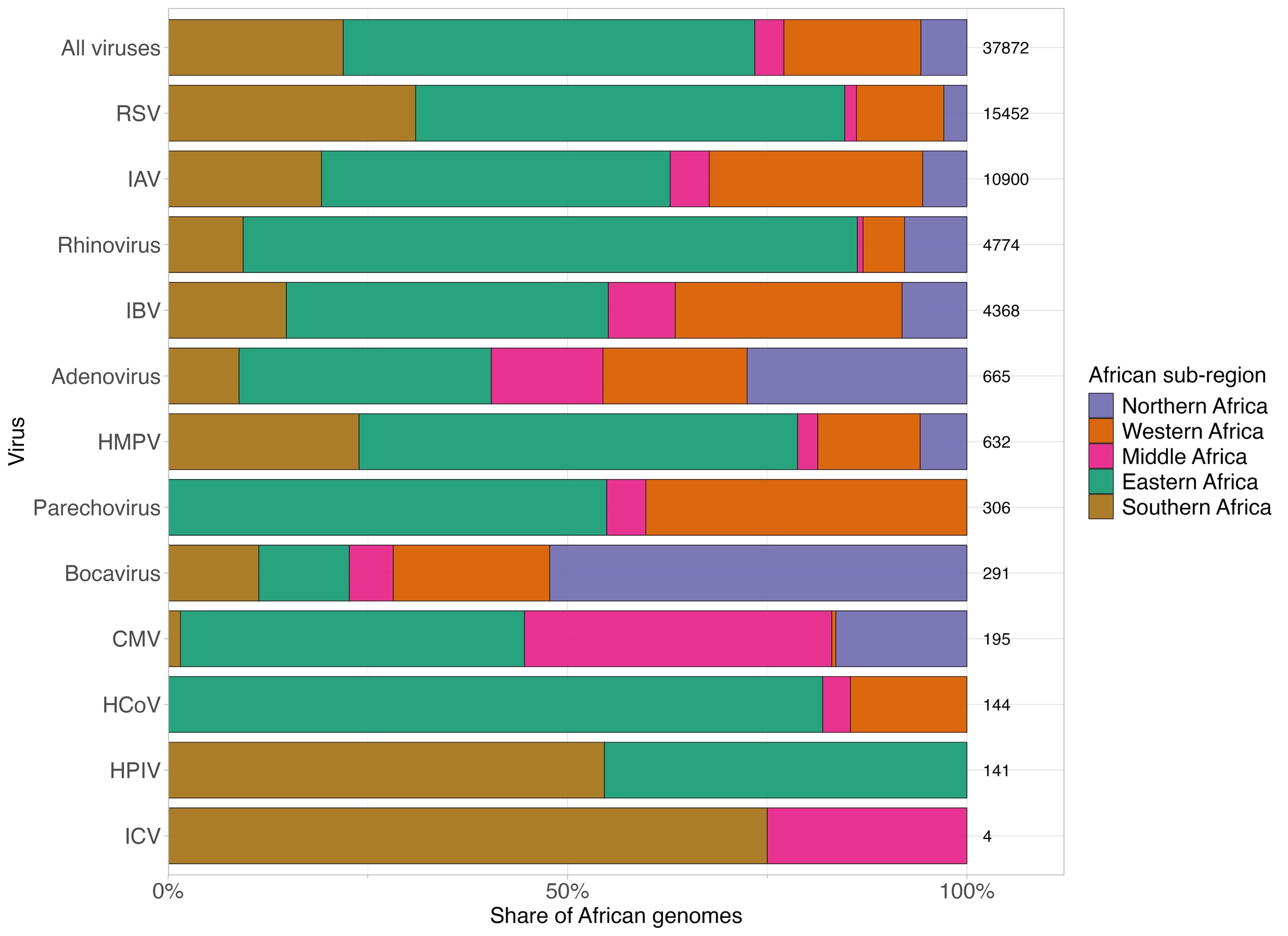

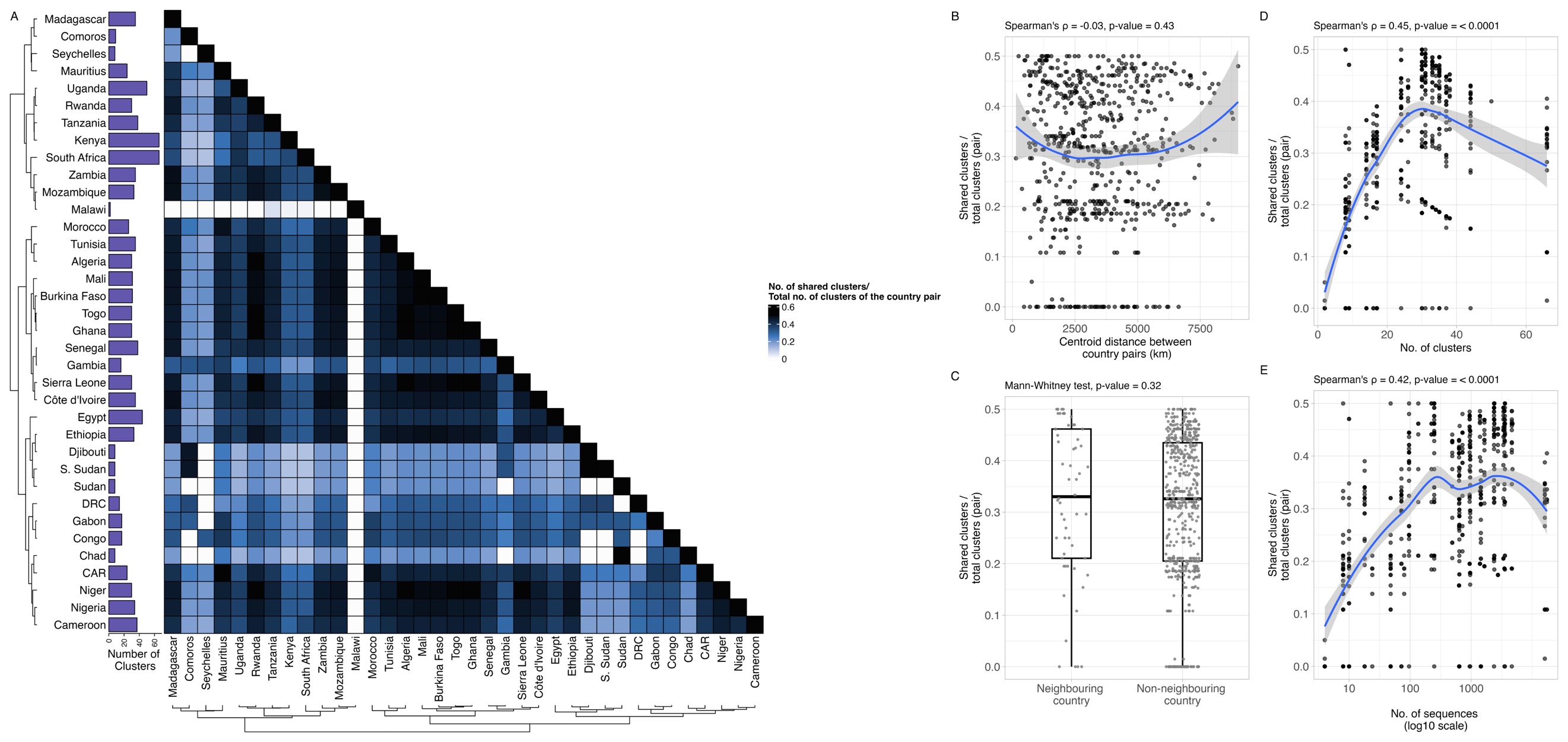

Supplementary Figure 2. Exploring patterns in cluster sharing to geographical proximity among African respiratory virus genomic sequences with ≥90% genome/segment coverage. (A) Heatmap of shared viral clusters between African countries. Colour denotes the proportion of shared clusters between two countries relative to the total number of clusters from both countries. The bar plot (left) indicates the number of clusters per country. The dendrogram shows the geographical proximity between countries, as inferred by the distance between the centroids of each country. Scatterplots shows the proportion of shared clusters between two countries relative to the total number of clusters from both countries against (B) the distance between the centroids of the two countries (Spearman’s test, p-value = 0·31), (D) the number of clusters (Spearman’s test, p-value < 0·0001), and (E) the number of sequences (Spearman’s test, p-value < 0·0001). LOESS smooth lines were shown to indicate general trends in panel B, D and E. (C) A boxplot compares the proportion of shared clusters between two countries relative to the total number of clusters from both countries for neighbouring and non-neighbouring countries, for which no significant difference was observed (Mann-Whitney test, p-value = 0·15). The central box denotes the interquartile range, while the horizontal zone within the box is the median.

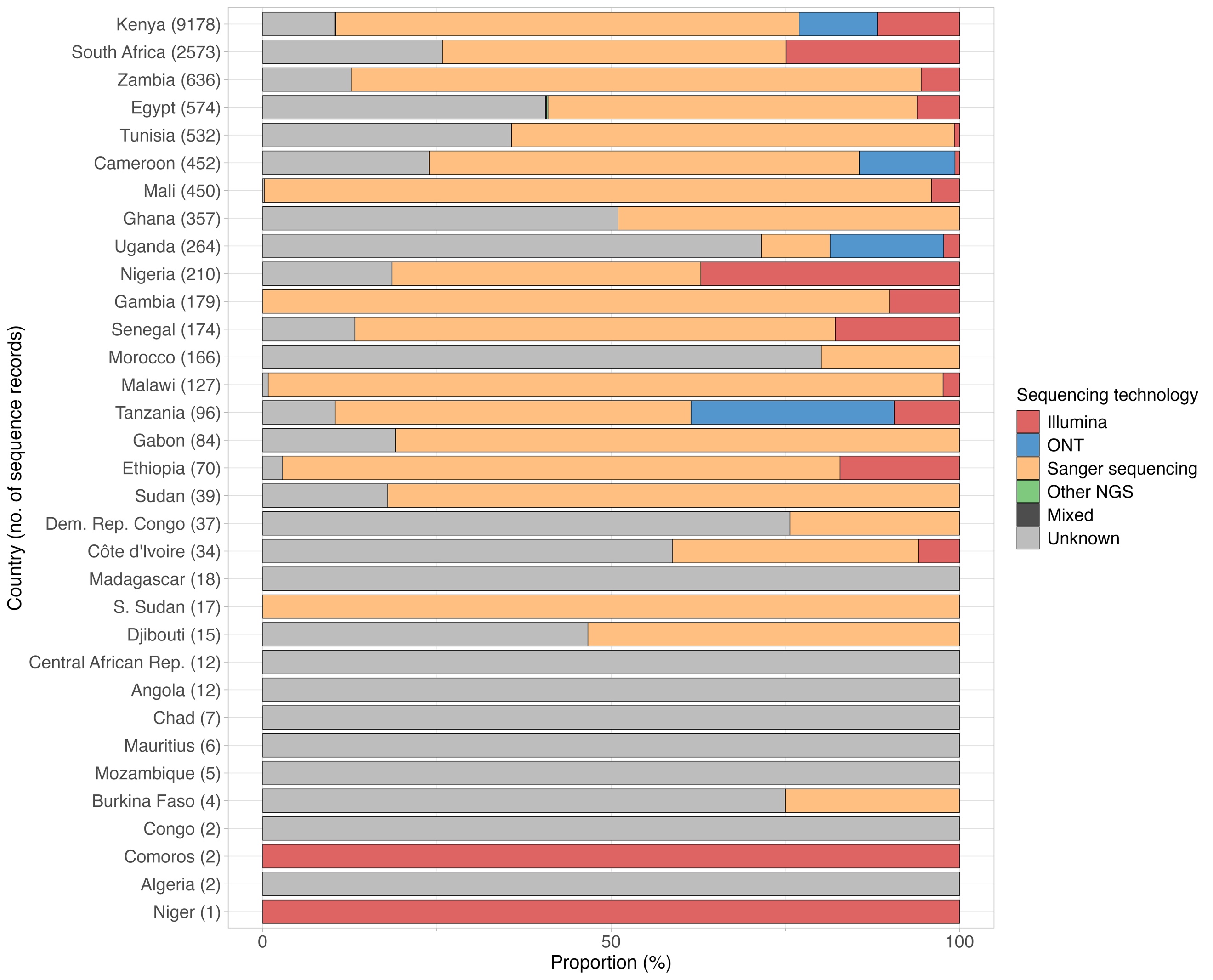

Supplementary Figure 3**.** Sequencing technology per African country/territory. Proportion of GenBank sequence records generated by each sequencing technology, for each country (total number of records in parentheses). Bars are coloured by sequencing technology; "Unknown" denotes records with no reported platform. All records from GISAID and Pathoplexus were excluded from this analysis due to insufficient metadata on the sequencing technology.

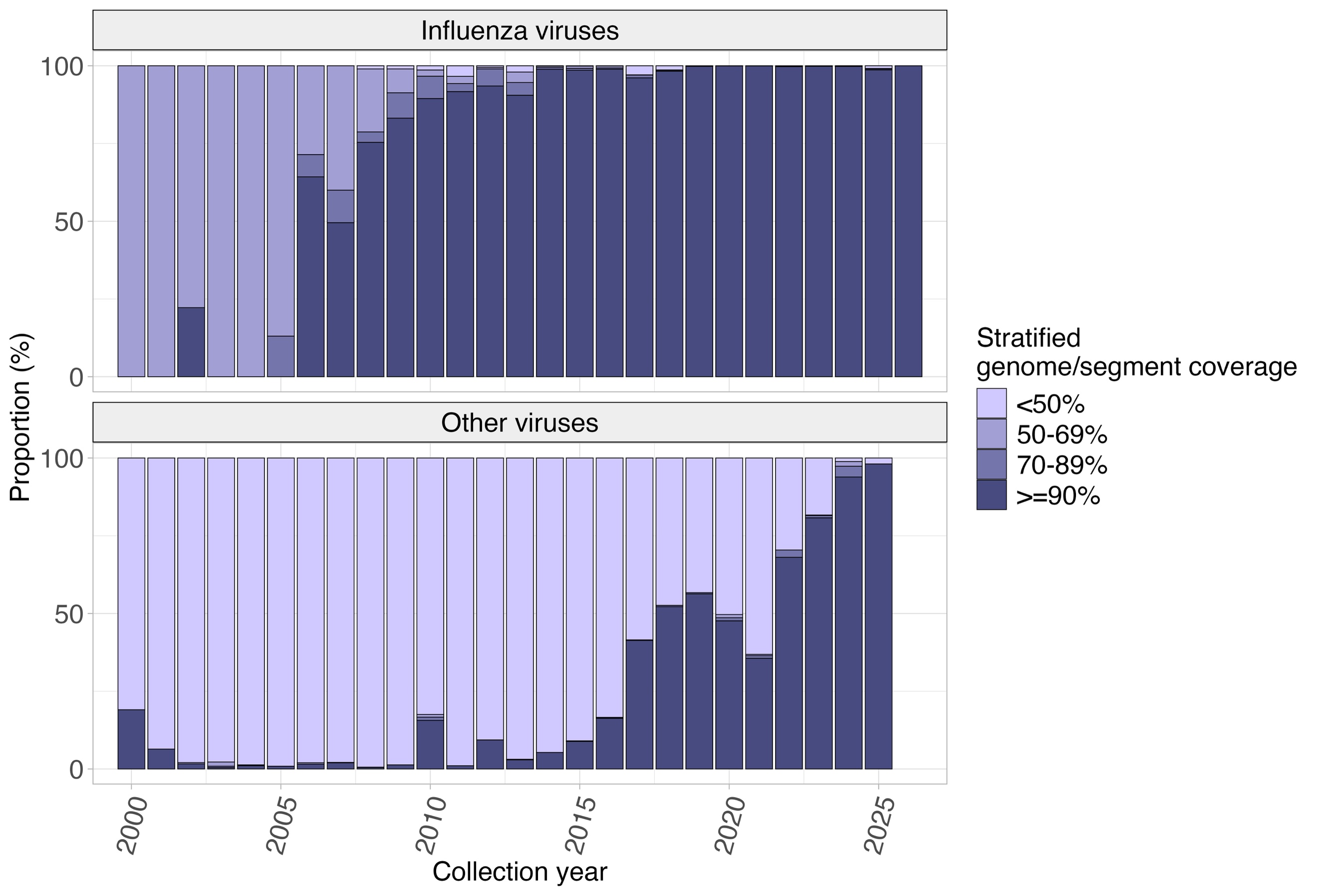

Supplementary Figure 4**.** Stratified genome/segment coverage of respiratory virus sequences from Africa over time. Bars show the proportion of sequences in each coverage band per collection year, with coverage assessed per segment for influenza A, B and C viruses (top) and per genome for all other viruses (bottom). Sequences with unspecified collection year were excluded from this analysis.

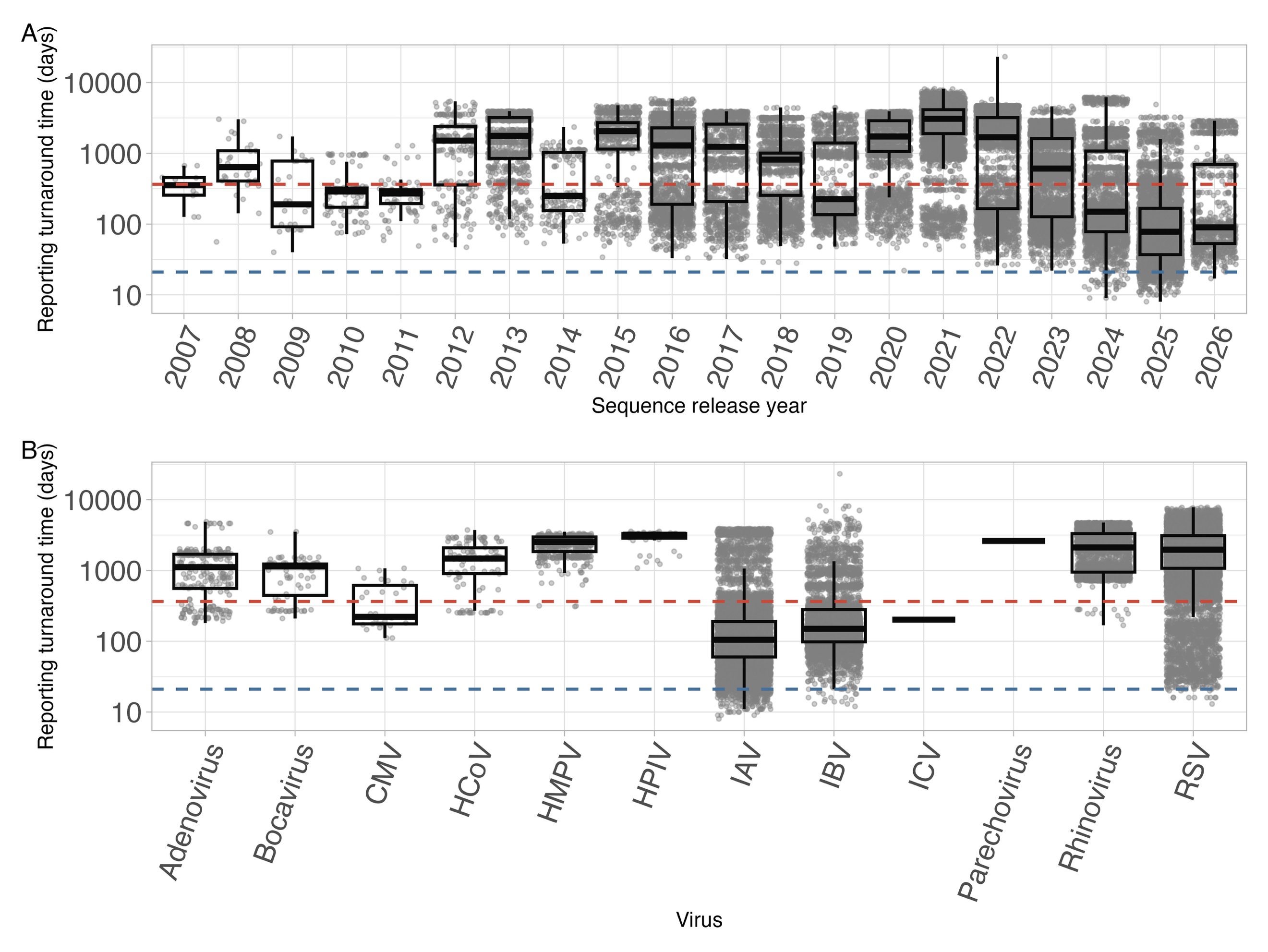

Supplementary Figure 5**.** Distribution of sequencing turnaround time of African respiratory virus sequences available per sequence release year (A) and per virus (B). Sequencing turnaround time was calculated by comparing the time difference between the sample collection and sequence release dates. The y-axis is on a log scale. Each dot represents a single deduplicated sequence record. The central box of each boxplot represents the interquartile range (IQR), with the horizontal line inside the box indicating the median. The blue horizontal line represents a rapid sequencing turnaround benchmark (21 days), while the red dashed line indicates a 365-day threshold. Sequence records with unspecified collection date were excluded from this analysis.

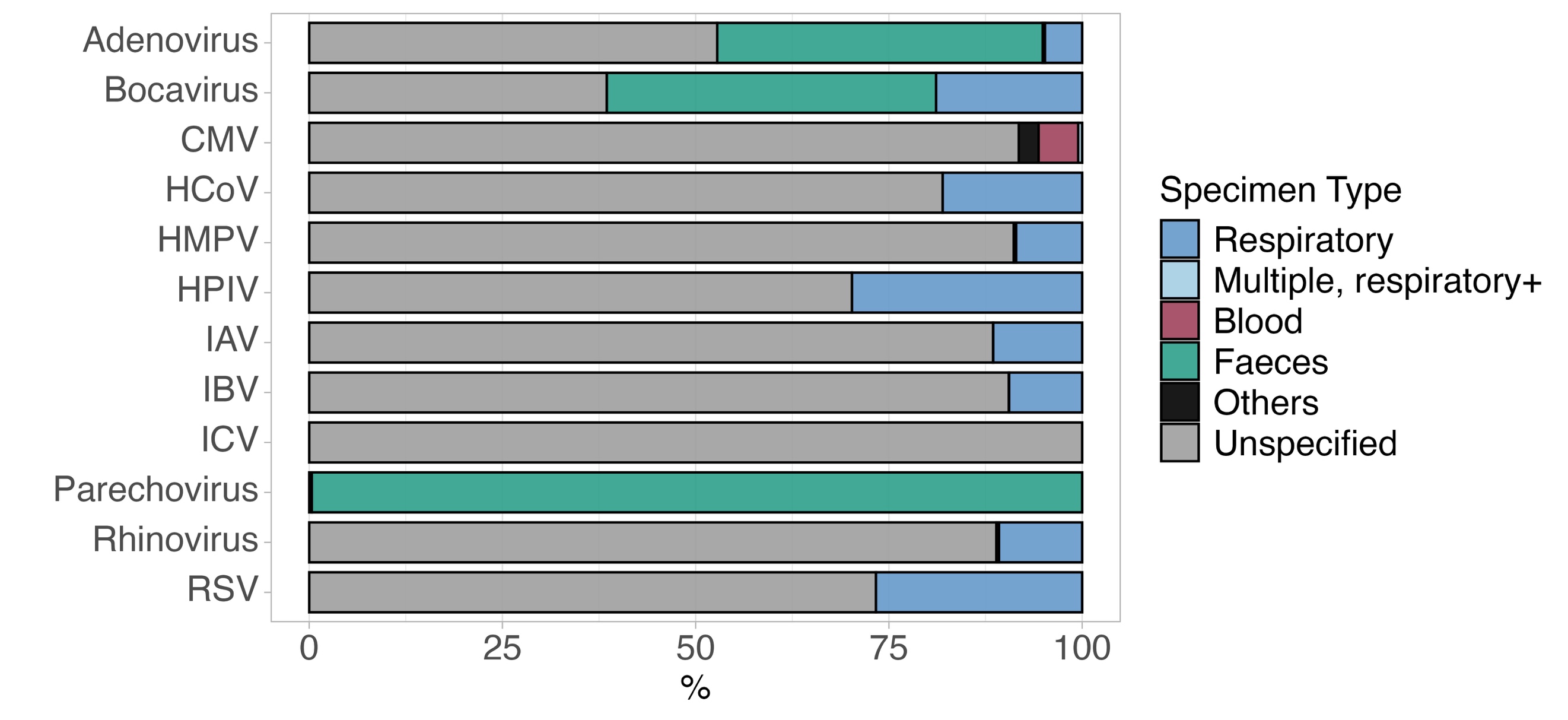

Supplementary Figure 6**.** Distribution of specimen type across respiratory virus sequences from Africa. The X-axis indicates the proportion of deduplicated sequence records. Bars are coloured by different specimen types. Records listing multiple specimen types that included at least one respiratory sample were classified as “Multiple, respiratory+”, while those listing multiple types excluding respiratory specimens were classified as “Multiple, respiratory-”. Specimens that could not be assigned to the primary categories were grouped under “others”.

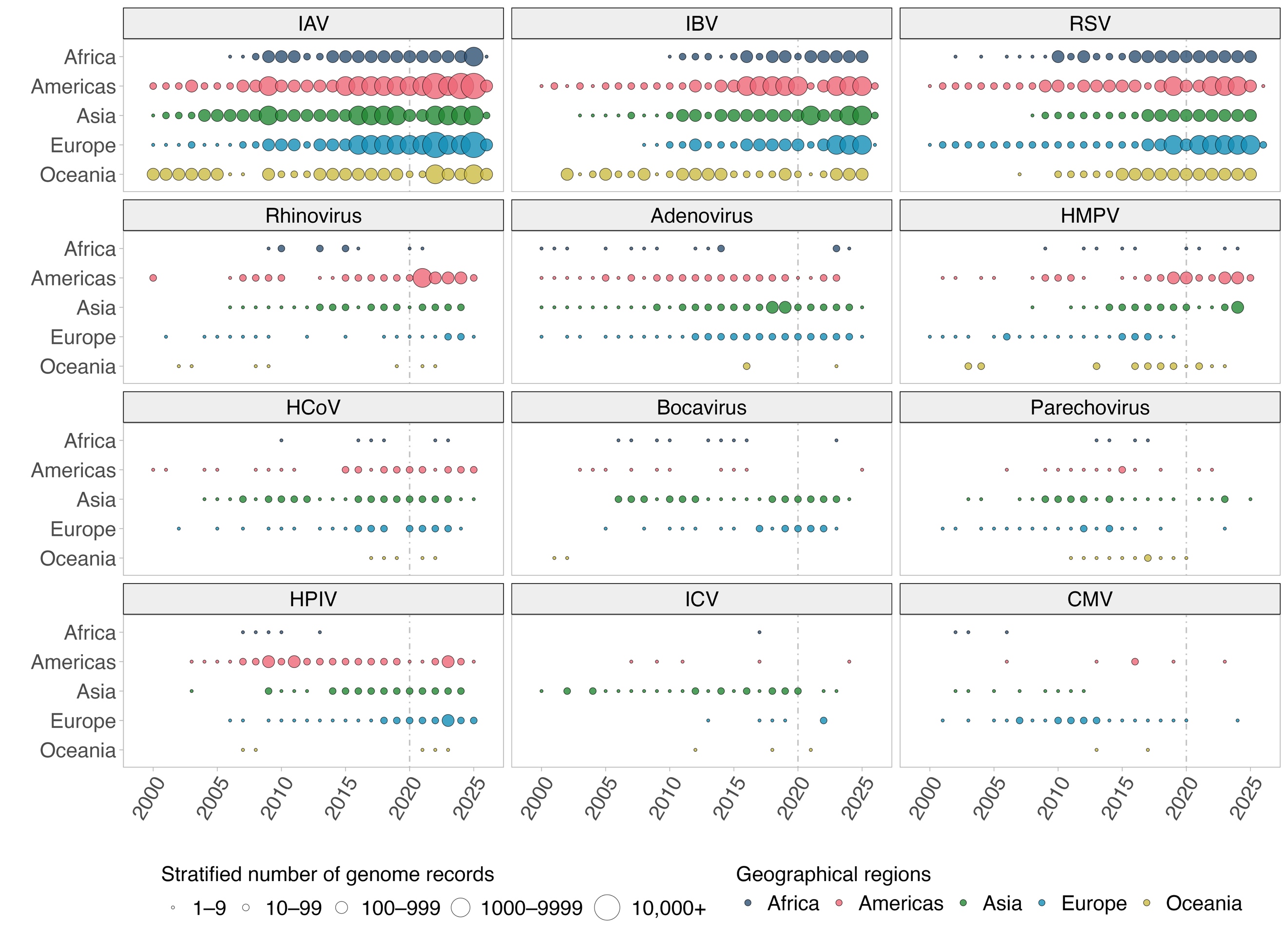

Supplementary Figure 7**.** Temporal distribution of deduplicated respiratory virus genomic sequence records across five geographical regions as defined by the United Nations Statistics Division, collected between 2000 and 2025. The colour of the bubbles denotes the geographical region, and the size of the bubbles indicates the stratified number of genome records. The grey vertical dotted line highlights the year 2020.

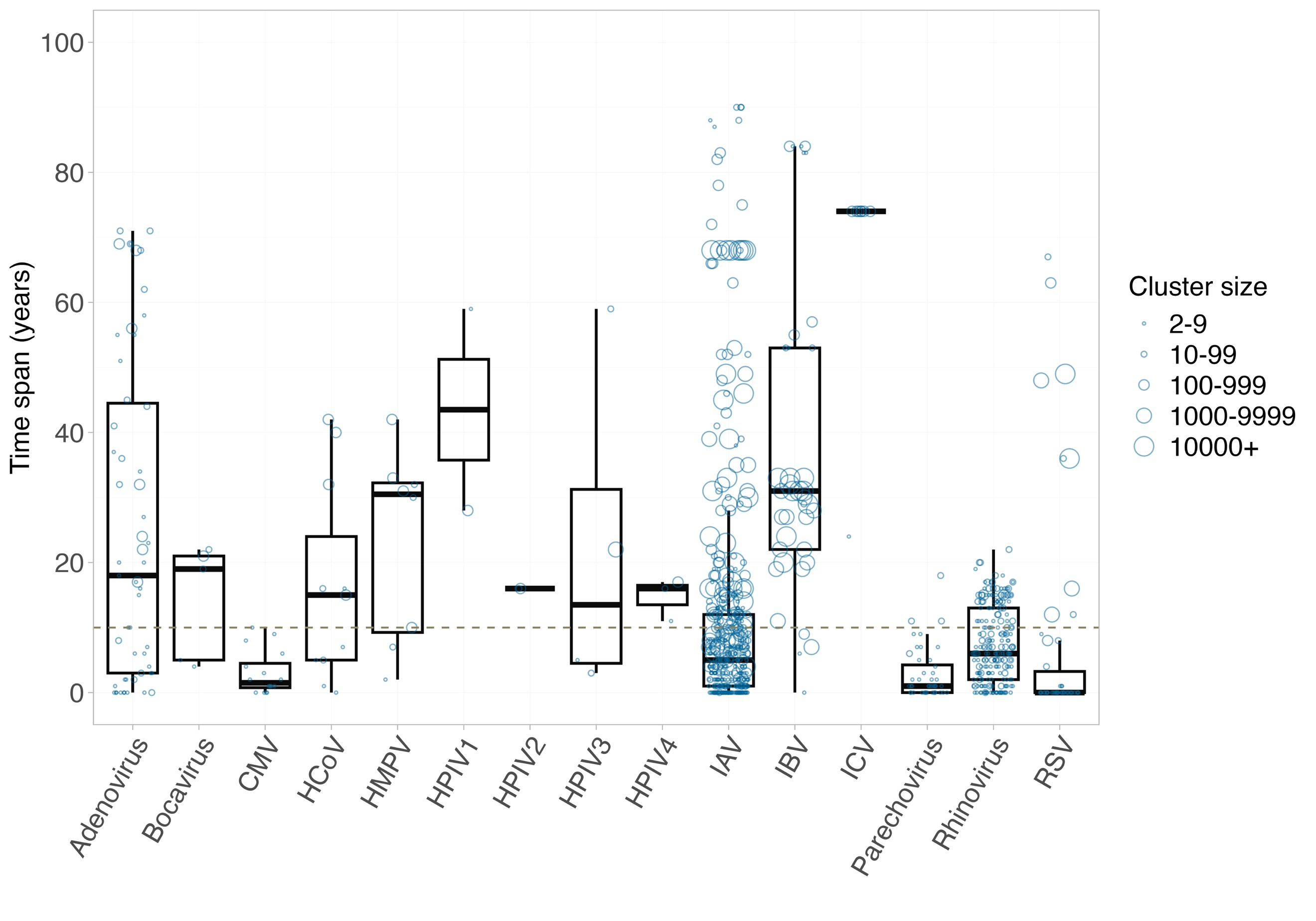

Supplementary Figure 8**.** Boxplot and jitter plot showing the time spans of non-singleton clusters of different respiratory viruses. Each circle represents one non-singleton cluster. The size of the circle denotes the stratified number of sequences (i.e. cluster size) of the cluster. The central box shows the interquartile range, while the horizontal line in the box represents the median. The khaki dashed line shows a 10-year time span.

### Supplementary Methods

#### Search strategy

##### GenBank

GenBank nucleotide sequence records of 12 study-defined groups of endemic human non-SARS-CoV-2 respiratory viruses were identified using their respective NCBI taxonomic identifiers (TaxId) via the NCBI Virus portal in March 2026 (**Supplementary Table** **1**). These viruses include influenza A (IAV), B (IBV) and C (ICV) viruses, respiratory syncytial virus (RSV), human parainfluenza virus 1–4 (HPIV), human coronaviruses HKU1, OC43, NL63 and 229E (HCoV), human metapneumovirus (HMPV), rhinovirus, parechovirus, adenovirus, bocavirus and cytomegalovirus (CMV). During the search process, filters were applied to include only sequences from human hosts (*Homo sapiens*, TaxId: 9606) and to exclude those labelled as originating from environmental sources, lab-passaged isolates or vaccine strains. Sequences containing more than 50 ambiguous nucleotides (i.e. N) were also excluded. No restrictions were imposed on the collection date, meaning that sequences without a collection date were included in our dataset.

##### GISAID

IBV and ICV sequences were downloaded from the GISAID EpiFlu™ database in June 2026, and RSV sequences were downloaded from EpiRSV™. The IBV and ICV searches were limited to human-derived sequences. IAV sequences were drawn from a previously curated dataset that already contains both GenBank and GISAID records. All sequences labelled with Temporary Publishing Embargo (TPE) were excluded.

##### Pathoplexus

All RSV-A, RSV-B and HMPV sequences were retrieved from Pathoplexus in March 2026. All sequences with restricted access were excluded.

#### Datasets

##### African dataset

All available partial and complete sequences collected from Africa were included in the African dataset. The country or territory of origin recorded for each sequence record was matched against a list of African countries defined by the United Nations Statistics Division (UNSD) Standard Country or Area Codes for Statistical Use. Records with geolocation could not be resolved to an UNSD African country/territory and were excluded.

##### Global genomic dataset

For the global genomic dataset, worldwide influenza A, B and C virus (IAV, IBV and ICV) genomes were retained only when all segments were present, each with ≥90% coverage, while all other (non-segmented) viruses were retained at ≥90% genome coverage to ensure sufficient completeness for reliable clustering. Genome coverage was estimated for all sequences based on their corresponding RefSeq complete genome sequences. Geographical regions were assigned using UNSD (M49) definitions; for locations not listed by UNSD, the region was assigned based on continent data from the rnaturalearth R package. Sequences without a region assignment were excluded. Population denominators used to express genome records per population for each world region were obtained from the 2024 Revision of the World Population Prospects (United Nations Department of Economic and Social Affairs, Population Division).

#### Data curation

##### Sequence validation

Sequences were validated by BLASTn against the corresponding NCBI RefSeq reference and reference genome sequences to confirm virus identity. Records that returned no hits were subjected to web-based BLASTn analysis against the NCBI nt database and excluded when identity could not be confirmed. Sequences with misassigned taxa or segments were assigned to the correct taxa/segments based on BLASTn results.

##### Eligibility filtering

A uniform set of eligibility criteria was applied to all repositories: only sequences from a human host, with ≤50 ambiguous bases (N) per sequence, were retained, and environmental, vaccine and laboratory-derived sequences were excluded. Additional keywords (i.e., vaccine, wild, clone, propagate, and FluMist) were used to filter sequences from experimental settings and/or vaccine strains. To ensure comparability across databases, a common cut-off of 10 February 2026 was applied, and only sequences released on or before this date were retained.

##### Record deduplication

To avoid redundant counting arising from repeated or cross-repository submissions, record deduplication was performed as follows. For non-segmented viruses, candidate duplicate records were identified within each virus by shared repository accession, by BioSample (for records lacking a usable strain name), or by a standardised strain name (derived from the strain, NCBI organism and isolate names). Candidate records were merged into a single representative only when their countries of origin were concordant; differences in collection dates were tolerated only when their nucleotide sequences were byte-identical (SHA-256 hash). Within each merged group, the longest sequence was retained as the representative, with repository priority GenBank > GISAID > Pathoplexus used to break ties of equal length. For segmented viruses (IAV, IBV, and ICV), records were grouped into a single genome per strain. Within each strain, a single representative sequence was retained per segment by selecting the longest sequence, with repository priority (GenBank > GISAID) used to break ties of equal length. Sequence-availability counts were therefore expressed as one record per strain for segmented viruses and one record per sequence for non-segmented viruses.

##### Spatial data

Spatial data, including geographical coordinates of each African country, was obtained from R package rnaturalearth. Shared border lengths and centroid distance between countries, used in the geographical proximity and cluster sharing frequency analyses, were estimated using the st_centroid and st_intersection functions from R package sf.
